# Virtual Recruitment is Here to Stay: 2020 ID Fellowship Program and Matched Applicant Recruitment Experiences

**DOI:** 10.1101/2021.05.07.21256828

**Authors:** Danica Rockney, Constance A. Benson, Brian G. Blackburn, Lisa M. Chirch, Victoria J. L. Konold, Vera P. Luther, Raymund R. Razonable, Sean Tackett, Michael T. Melia

## Abstract

**Background:** Graduate Medical Education training programs transitioned to all-virtual recruitment in 2020. Few data have been published regarding the consequences of this transition. We desired to understand (1) infectious diseases (ID) fellowship programs’ recruitment efforts and the effect of virtual recruitment on application and interview numbers, and (2) the number of programs to which matched applicants applied and interviewed, and their perspectives on virtual recruitment.

**Methods:** In 2020-21 we surveyed all United States ID fellowship program directors (PDs) and matched applicants. Descriptive data analysis was performed on quantitative survey items. Free-text responses were analyzed through a quantitative content analysis approach.

**Results:** PD response rate was 68/158 (43%); applicant response rate was at least 23% (85/365). PDs reported a 27% increase in mean number of applications received and a 45% increase in mean number of applicants interviewed. Applicants especially valued online program structure information, PD program overview videos, fellow testimonials, didactic and curriculum content, and current fellow profiles. Most applicants preferred interviews lasting no more than 40 minutes and interview days lasting no more than 5 hours. Nearly all (60/64, 94%) PDs adequately learned about candidates; most (48/64, 75%) felt unable to showcase their program as well as when in-person. Most PDs (54/64, 84%) and applicants (56/73, 77%) want at least an option for virtual recruitment moving forward.

**Conclusions:** Virtual recruitment enabled programs to accommodate more applicants and highlighted applicants’ preferences for programs’ augmented online presences and time-limited interview days. Most programs and applicants want the option for virtual interviews.

**Main Points:** Virtual recruitment enables programs to accommodate more applicants. Applicants value programs’ augmented online presences and favor time-limited interview days. Most programs and applicants prefer in-person interviews and want at least an option for virtual interviews.

## Introduction

Because of the COVID-19 pandemic, for the 2020-2021 recruitment season the Coalition for Physician Accountability recommended all residency and fellowship programs “commit to online interviews and virtual visits for all applicants, rather than in-person interviews for the entire [2020-21] cycle.”^1^ As a result, programs were compelled to establish virtual interviewing processes and platforms while concomitantly trying to upgrade their online presences. Meanwhile, applicants needed to adjust their expectations for interviewing and learning about new programs and cities from afar.

To guide applicants and programs, a number of perspective pieces, editorials, viewpoints, reviews, and single-program experiences were published prior to, during, and after the medical specialty fellowship program recruitment season;^2-19^ one review provided evidence-based best practices.^20^ Other contributions discussed the importance of mindfulness regarding potential biases against applicants under-represented in medicine and the potential role of social media.^4,21-23^

Prior specialty-wide study of virtual recruitment has largely been limited to surgical specialties, many of which transitioned from in-person to virtual recruitment in the middle of their 2020 recruitment seasons. In a survey of complex general surgical oncology program directors (PDs) and applicants, most PDs felt virtual interviews permitted accurate portrayals of programs and applicants.^24^ Roughly half of applicants felt virtual interviews allowed applicants to accurately portray themselves; nearly half had a neutral view regarding programs’ ability to accurately represent themselves. In another report, over 85% of surveyed female pelvic medicine and reconstructive surgery fellowship PDs reported satisfaction with virtual interviews and found them effective in assessing applicants; 31% preferred virtual interviews, and 60% reported being likely to offer virtual interviews in the future.^25^ Surveys of applicants to and faculty of other programs found that applicants and faculty preferred in-person interviews, felt they did not get to know one another as well virtually as in-person, and were less able to understand program culture and make an informed rank list.^26-28^ To our knowledge, there are no published data regarding the efforts undertaken by programs in advance of an entirely virtual recruitment season, the effect of all-virtual recruitment on application numbers, or the perspectives of PDs and applicants on all-virtual recruitment within non-surgical specialties.

Through a survey of infectious diseases (ID) fellowship PDs we aimed to understand the impact of virtual recruitment on the number of applications received by programs, the number of interviews offered and conducted, and the recruitment resources developed by programs. Through a survey of matched applicants to these programs, we aimed to understand the number of programs to which applicants applied and interviewed and their perspectives on discrete components of virtual recruitment.

## Methods

Authors drafted and revised survey questions. Survey items were finalized through consensus.

On December 14, 2020 one author (MTM) sent emails to each ID fellowship PD requesting their participation in the PD survey (Supplementary Appendix 1). These emails were sent 12 days after Match day and contained a hyperlink to the survey.

Because there is no central repository containing the names and contact information for all applicants who matched into ID fellowship programs, we asked each PD to email the applicant survey request to applicants who matched into their program (Supplementary Appendix 2). PDs were sent a draft email to be sent to each matched fellow requesting their participation in the survey. This draft email contained a hyperlink to the applicant survey. We asked applicants to reflect not only on their experiences with the program with which they matched, but upon their collective recruitment experiences. We were not able to contact unmatched applicants.

These emails were sent once weekly for four weeks. To provide reminders and help ensure receipt of our messages, the day after each email was sent one author (MTM) sent a message through the Training PD Community (i.e., listserv) within the online MyIDSA platform of the Infectious Diseases Society of America (IDSA).

All responses were anonymous. In order to protect and maintain participant anonymity, we did not solicit demographic data about program directors, programs, or applicants.

Descriptive data analysis was performed on quantitative survey items. Questions that asked for free-text input of numerical data occasionally solicited impossible values (e.g., >100% of applicants were interviewed). These values were excluded from analysis as noted in each table. We used t-tests to compare differences in mean applications, interview invitations, and interviews comparing 2020 to 2019 in the program director surveys. Analyses were performed using Stata (StataCorp. 2013. *Stata Statistical Software: Release 13*. College Station, Texas: StataCorp LP.) Because responses to open-ended survey prompts are typically not appropriate for formal qualitative approaches to analysis, free-text responses to questions regarding aspects of virtual recruitment to retain or change (questions 19 and 20 on each survey) were analyzed through a quantitative content analysis approach.^29^ A single author (MTM) reviewed all responses and coded them. These codes and frequencies were reviewed by an additional author (DR) and discrepancies were resolved through discussion.

### Conflicts of Interest

None of the authors has any relevant conflicts of interest.

### Patient Consent Statement

This study was acknowledged as exempt by the Johns Hopkins University School of Medicine Institutional Review Board. This study does not include factors necessitating patient consent.

## Results

### Program Director Survey

The survey was sent to all 158 United States adult ID fellowship PDs. Sixty-eight (43%) responded.

PDs reported a 27% increase in the mean number of applications submitted to their programs [89.2 (SD 50.6) vs 70.2 (SD 48.3), p=.03], a 23% increase in the mean number of interview invitations offered [41.7 (SD 16.9) vs 34.0 (SD 14.4), p<0.01], and a 45% increase in the mean number of applicants interviewed [34.9 (SD 17.0) vs 24.0 (13.6), p<0.01] in 2020 as compared with 2019 (Table 1a). While there was no significant change in the proportion of applicants invited to interview, 81% of applicants who were offered an interview in 2020 attended an interview day, up from 68% in 2019 (p<0.01). The majority (48/67, 72%) of PDs anticipate interviewing the same number of applicants in 2021 as in 2020, with only 8 (12%) and 11 (16%) planning to interview fewer or more applicants, respectively.

**Table 1a:**
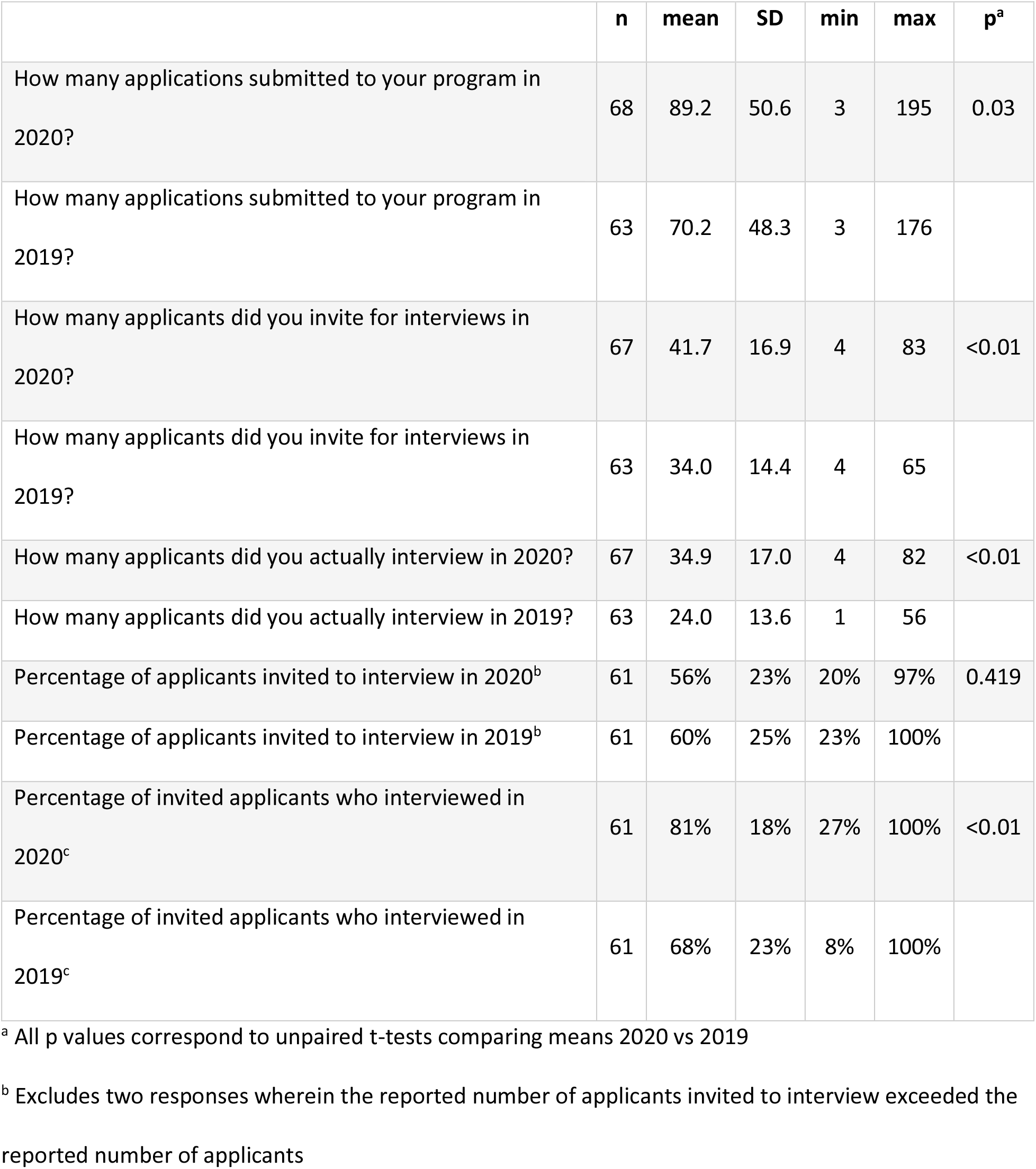
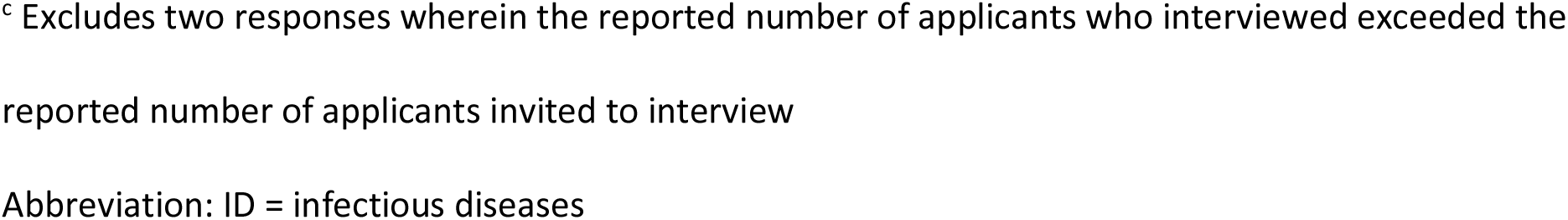
Application and Interview Data Reported by ID Fellowship Program Directors for the 2020 Recruitment Season

|  | n | mean | SD | min | max | p<sup>a</sup> |
| --- | --- | --- | --- | --- | --- | --- |
| How many applications submitted to your program in 2020? | 68 | 89.2 | 50.6 | 3 | 195 | 0.03 |
| How many applications submitted to your program in 2019? | 63 | 70.2 | 48.3 | 3 | 176 |  |
| How many applicants did you invite for interviews in 2020? | 67 | 41.7 | 16.9 | 4 | 83 | <0.01 |
| How many applicants did you invite for interviews in 2019? | 63 | 34.0 | 14.4 | 4 | 65 |  |
| How many applicants did you actually interview in 2020? | 67 | 34.9 | 17.0 | 4 | 82 | <0.01 |
| How many applicants did you actually interview in 2019? | 63 | 24.0 | 13.6 | 1 | 56 |  |
| Percentage of applicants invited to interview in 2020<sup>b</sup> | 61 | 56% | 23% | 20% | 97% | 0.419 |
| Percentage of applicants invited to interview in 2019<sup>b</sup> | 61 | 60% | 25% | 23% | 100% |  |
| Percentage of invited applicants who interviewed in 2020<sup>c</sup> | 61 | 81% | 18% | 27% | 100% | <0.01 |
| Percentage of invited applicants who interviewed in 2019<sup>c</sup> | 61 | 68% | 23% | 8% | 100% |  |

<sup>a</sup> All p values correspond to unpaired t-tests comparing means 2020 vs 2019
<sup>b</sup> Excludes two responses wherein the reported number of applicants invited to interview exceeded the reported number of applicants
<sup>c</sup> Excludes two responses wherein the reported number of applicants who interviewed exceeded the reported number of applicants invited to interview
Abbreviation: ID = infectious diseases

**Table 1b:** Application and Interview Data Reported by Matched ID Fellowship Program Applicants for the 2020 Recruitment Season

|  | **n** | **mean** | **SD** | **min** | **max** |
| --- | --- | --- | --- | --- | --- |
| To how many programs did you apply in 2020? | 82 | 20.2 | 15.3 | 1 | 91 |
| How many interview offers did you receive in 2020? | 82 | 13.9 | 9.7 | 1 | 56 |
| How many interview days did you actually attend in 2020?<sup>a</sup> | 81 | 10.9 | 5.9 | 1 | 30 |

<sup>a</sup> One response excluded (respondent reported 0 interviews attended)
Abbreviation: ID = infectious diseases

The proportion of programs that generated, modified, or maintained without change different recruitment-related content is detailed in Table 2. The majority of programs newly created a program overview video, links to virtual tour(s), instructions on the technological aspects of virtual interviewing, presentations from fellows, and presentations from non-PD faculty.

**Table 2a:** Information made available by programs to applicants*

|  | No<sup>a</sup> |  | Yes,<br>maintained<sup>b</sup> |  | Yes,<br>modified<sup>c</sup> |  | Yes,<br>new<sup>d</sup> |  | Total |  |
| --- | --- | --- | --- | --- | --- | --- | --- | --- | --- | --- |
|  | n | % | n | % | n | % | n | % | n | % |
| Professionally- or program-made<br>program overview video | 18 | 26.9 | 2 | 3.0 | 2 | 3.0 | 45 | 67.2 | 67 | 100 |
| Program overview video by PD covering<br>program nuts and bolts | 30 | 45.5 | 0 | 0.0 | 10 | 15.2 | 26 | 39.4 | 66 | 100 |
| Presentations/testimonials from faculty<br>other than PD | 29 | 43.3 | 2 | 3.0 | 2 | 3.0 | 34 | 50.8 | 67 | 100 |
| Presentations/testimonials from fellows | 20 | 29.9 | 4 | 6.0 | 5 | 7.5 | 38 | 56.7 | 67 | 100 |
| Presentations/testimonials from<br>program alumni | 47 | 71.2 | 4 | 6.1 | 1 | 1.5 | 14 | 21.2 | 66 | 100 |
| Live, virtual attendance at<br>departmental/division conference(s) | 27 | 40.3 | 6 | 9.0 | 8 | 11.9 | 26 | 38.8 | 67 | 100 |
| Links to departmental/division<br>conference(s) to permit asynchronous<br>attendance | 44 | 66.7 | 1 | 1.5 | 1 | 1.5 | 20 | 30.3 | 66 | 100 |
| Profiles of current fellows | 14 | 21.2 | 21 | 31.8 | 17 | 25.8 | 14 | 21.2 | 66 | 100 |
| List of alumni and where they went<br>after fellowship graduation | 18 | 26.9 | 20 | 29.9 | 16 | 23.9 | 13 | 19.4 | 67 | 100 |
| Prose and photo-based content on program structure, training sites, clinical rotations, and research requirements | 4 | 6.0 | 16 | 23.9 | 26 | 38.8 | 21 | 31.3 | 67 | 100 |
| Prose and photo-based content on didactics and curriculum | 7 | 10.5 | 19 | 28.4 | 26 | 38.8 | 15 | 22.4 | 67 | 100 |
| Links to content from institutional GME office | 12 | 17.9 | 22 | 32.8 | 13 | 19.4 | 20 | 29.9 | 67 | 100 |
| Links to virtual tour(s) of your hospitals, clinic sites, university, campus, and/or clinical setting | 22 | 32.8 | 3 | 4.5 | 2 | 3.0 | 40 | 59.7 | 67 | 100 |
| Instructions for technological aspects of the interview season (e.g., description of format, computer/laptop, internet access, virtual platform, or other information) | 21 | 31.3 | 1 | 1.5 | 3 | 4.5 | 42 | 62.7 | 67 | 100 |
| Other | 2 | 25.0 | 0 | 0.0 | 1 | 12.5 | 5 | 62.5 | 8 | 100 |

\* on either a public-facing website as of 8/12/20 (the date on which applications became available to PDs) or otherwise
<sup>a</sup> No, we did not incorporate this content
<sup>b</sup> Yes; we maintained previously-created content without change
<sup>c</sup> Yes; we modified previously-created content for 2020 recruitment
<sup>d</sup> Yes; we newly created this content for 2020 recruitment
Abbreviations: PD = program director; GME = graduate medical education

**Table 2b:**
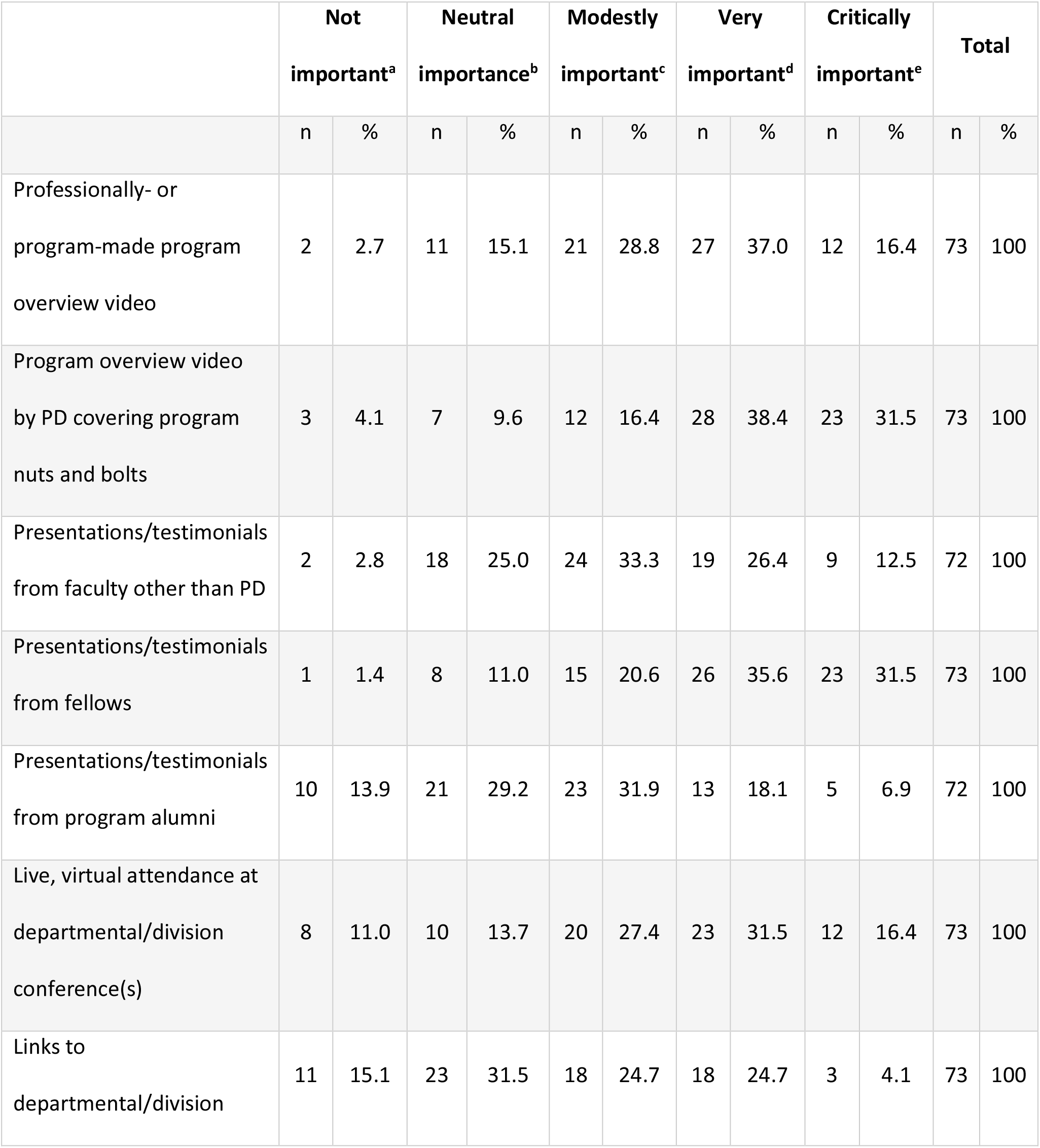
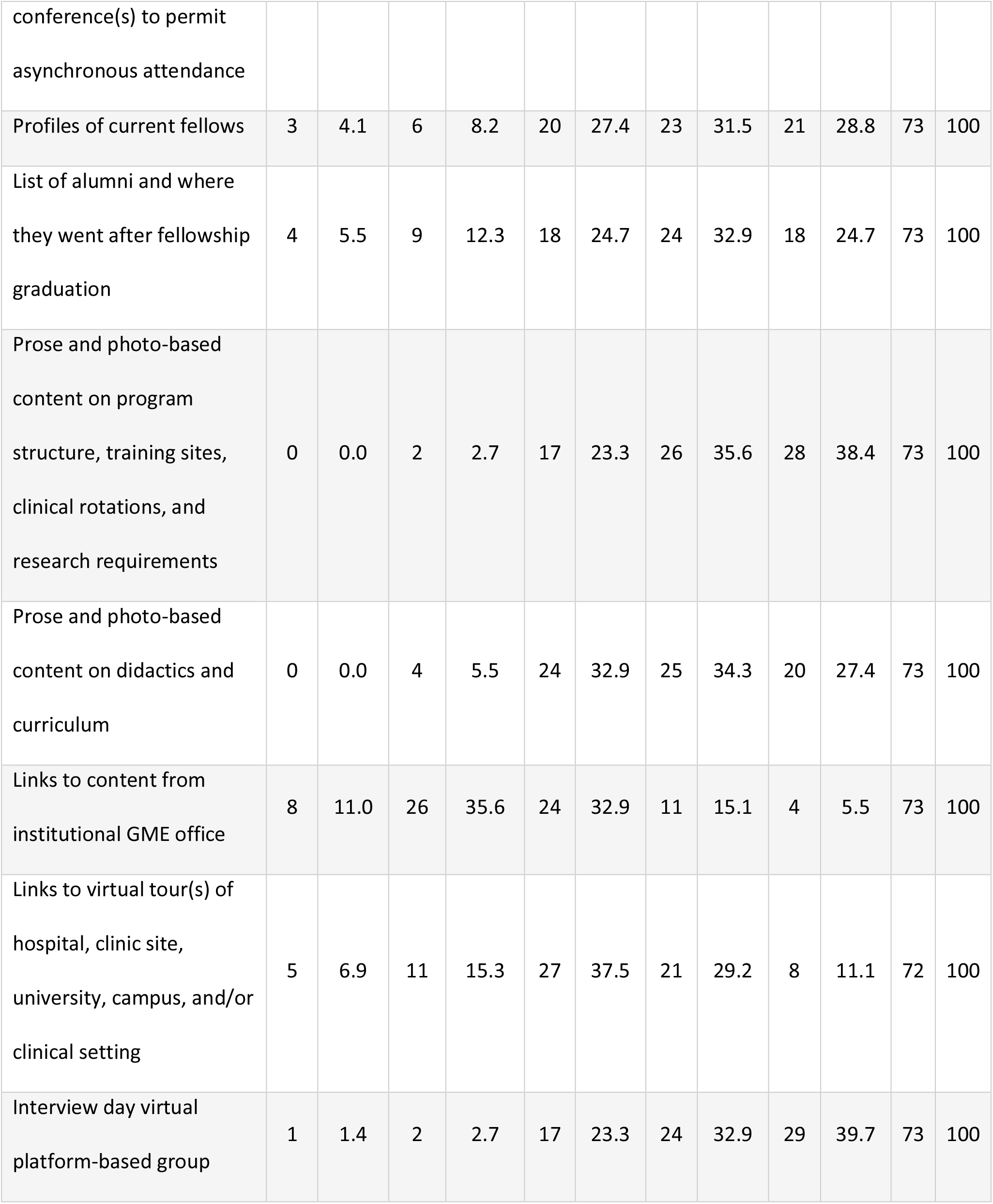
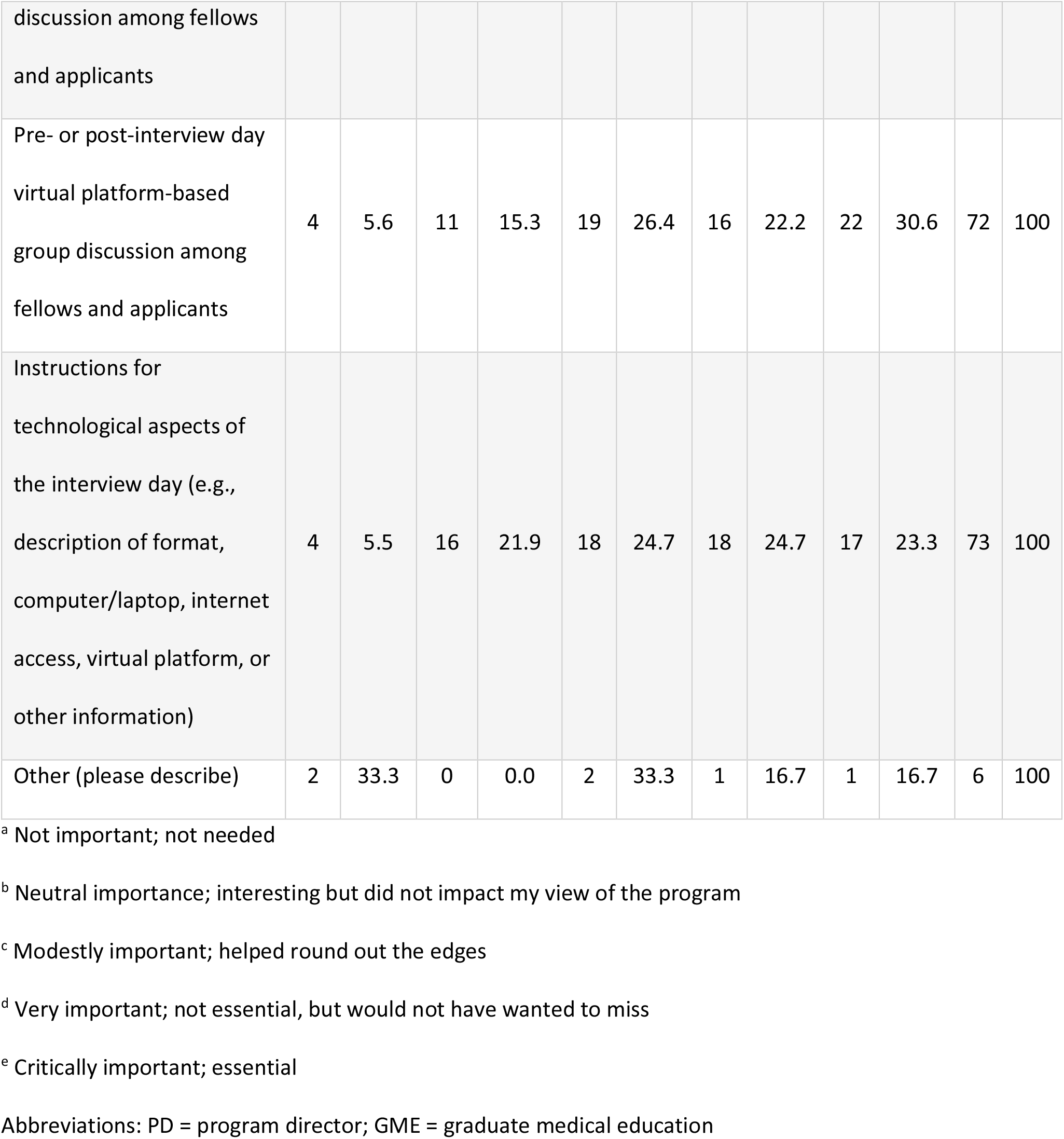
Matched applicants’ views of different features of programs’ websites

Most (57/67, 85%) programs required additional resources to facilitate the transition to a virtual format. For these programs, the amount of additional faculty and staff time varied. The majority of programs utilized at least 20 additional hours of PD + associate PD time, and the majority of programs required 10-29 additional hours of staff/personnel time (Table 3). The majority (28/55, 51%) of programs did not incur additional monetary expenses; these costs ranged from no additional monetary cost to a maximum of $25,000 for one program (Table 4). Half (32/64) of respondents said they will require fewer resources to support virtual recruitment should it be needed in 2021, whereas 26 (41%) will need similar resources and 6 (9%) more resources.

**Table 3.**
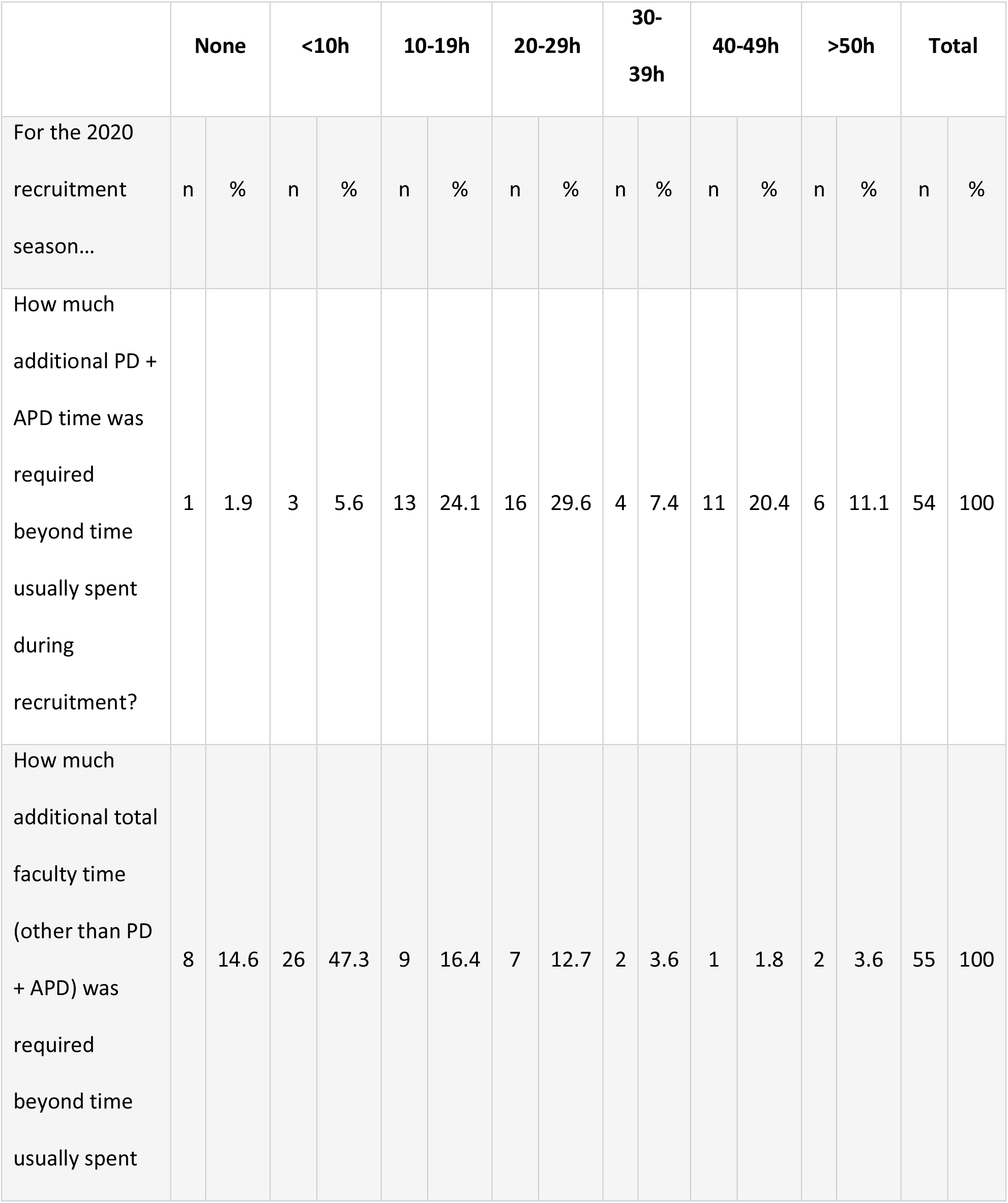
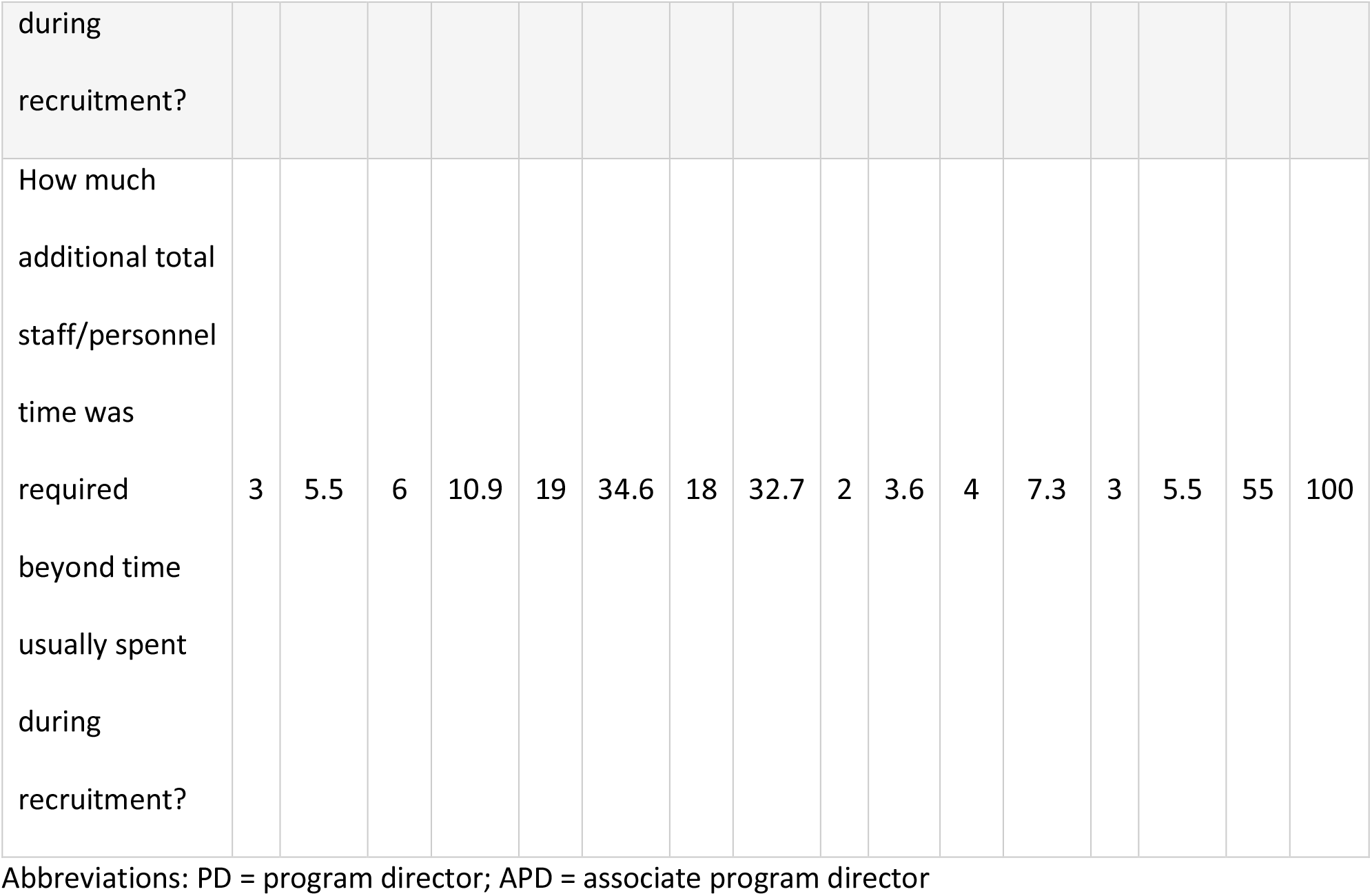
Self-reported additional time spent on virtual recruitment by program faculty and staff

**Table 4:**
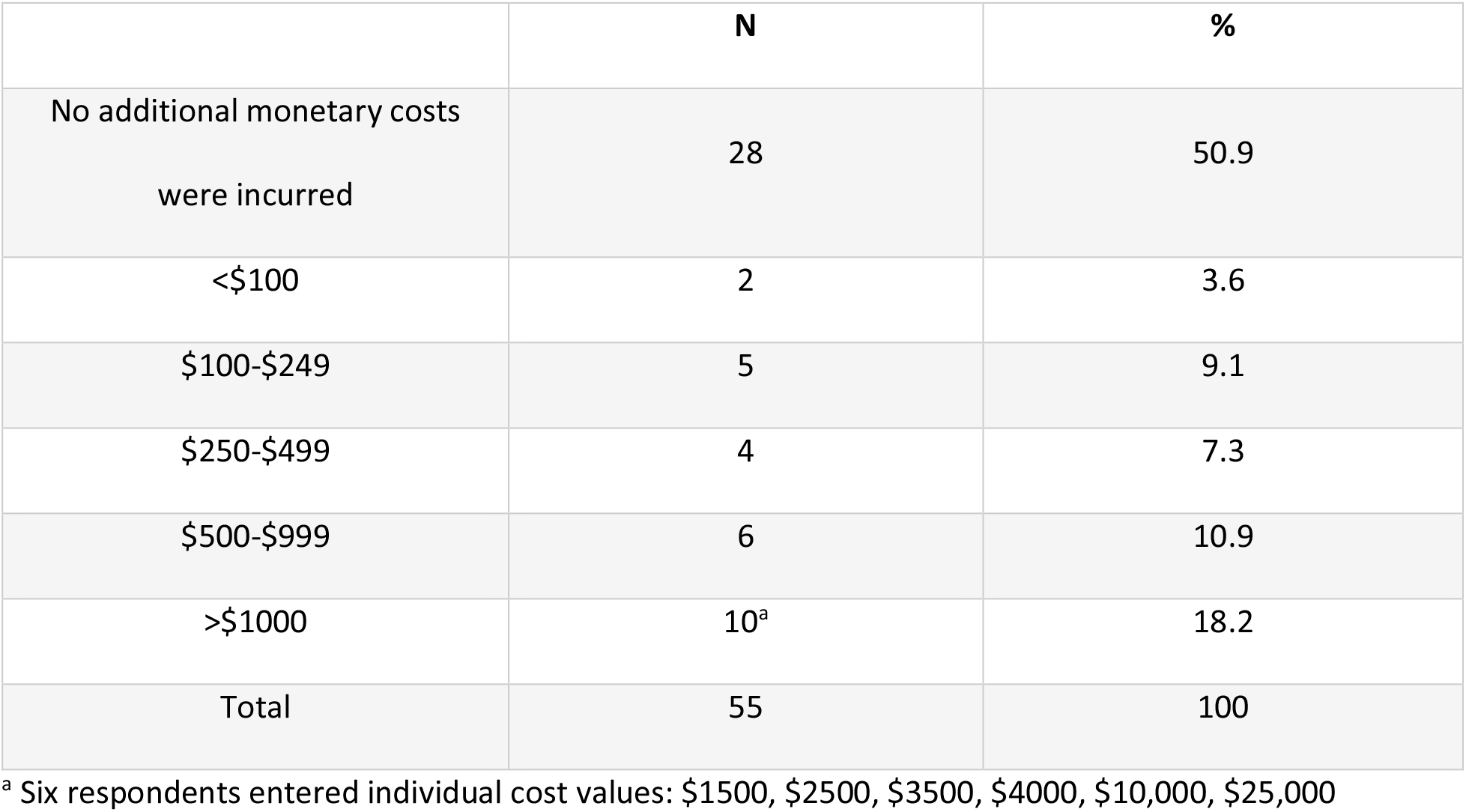
Self-reported monetary cost of virtual recruitment to programs

Nearly all (60/64, 94%) PDs felt they were able to sufficiently learn about each candidate via virtual recruitment, with 25 (39%) feeling they learned about applicants sufficiently but not as well as in-person, 31 (48%) equally well as in-person, and 4 (6%) better than in-person. Most PDs (48/64, 75%), however, felt they were either unable to adequately showcase their program (8, 12%) or were able to showcase their program adequately but less well than with in-person recruitment (40, 62%) Only 12 (19%) PDs felt they were able to showcase their program as well as in-person, and 4 (6%) better than in-person. Despite these concerns, most (54/64, 84%) PDs want to at least have the option for virtual recruitment moving forward, with 37 (58%) preferring face-to-face with an option for virtual, 9 (14%) preferring virtual with an option for face-to-face, and 8 (12%) preferring virtual. Only 10 (16%) prefer in-person interviewing with no virtual option.

### Matched Applicant Survey

There were 85 unique responses from matched applicants. The number of matched applicants who were sent the survey is not known; 365 positions were filled through the Match, so the response rate was at least 23% (85/365). These matched applicants applied to a mean of 20 programs (Table 1b). For 47/85 (55%) applicants, this number was not affected by the virtual nature of the 2020 recruitment season. Of the 38 for whom this number was affected, 13 (34%) applied to 1-3 additional programs, 18 (47%) applied to 4 or more additional programs, 4 (11%) applied to fewer programs, and 3 (8%) did not answer. In retrospect, the majority (52/73, 71%) of respondents would have applied to the same number of programs were they to repeat the experience, 19 (26%) would have applied to fewer programs, and few (2/73, 3%) would have applied to more programs.

Matched applicants received a mean of 14 interview offers and attended a mean of 11 interview days (Table 1b). Of the 48/82 (58%) of applicants whose decision to interview at programs was affected by the pandemic, 40 responded to a question quantifying this effect; half (20/40) of these respondents reported interviewing at 1-3 more programs than they otherwise would have, and half (20/40) at 4 or more. In retrospect, the majority (47/73, 64%) of respondents would have interviewed at the same number of programs were they to repeat the experience, and 26 (36%) would have interviewed at fewer; none would have interviewed at more programs.

The following components of the information made available by programs were deemed either critically or very important by at least 60% of applicants: prose- and photo-based content on program structure (54/73, 74%), group discussions with fellows (53/73, 73%), program overview video by PD (51/73, 70%), presentations from fellows (49/73, 67%), prose- and photo-based content on didactics and curriculum (45/73, 62%), and profiles of current fellows (44/73, 60%) (Table 2b). Three items were rated as of either neutral importance or unimportant by at least 40% of respondents: links permitting asynchronous conference attendance (34/73, 46%), links to institutional GME office content (34/73, 46%), and presentations from program alumni (31/72, 43%).

There was near-consensus regarding the structure of the interview day, with 84% (61/73) of matched applicants favoring 3-4 faculty interviews per day (Table 5). Nearly all (70/73, 96%) prefer interviews lasting less than 40 minutes, with 47/73 (64%) favoring interviews less than 30 minutes’ duration. The majority (48/73, 66%) of applicants preferred 5-9 minutes between interviews, with 13 (18%) preferring less than 5 minutes and 12 (16%) 10-14 minutes. Most (56/73, 77%) matched applicants prefer an interview day spanning 3-5 hours, and nearly all (65/73, 89%) prefer a single, consolidated interview day. Only 3/72 (4%) of applicants reported changing their rank order list based upon gift boxes or vouchers for food provided by programs; 31 (43%) viewed these favorably but did not change their rank order list as a result, 36 (50%) had a neutral view, and 2 (3%) an unfavorable view.

**Table 5:** Matched applicant perspectives on the structure and duration of the interview day

| Ideal number interviews/d |  |  | Ideal interview duration |  |  | Ideal virtual interview day duration |  |  |
| --- | --- | --- | --- | --- | --- | --- | --- | --- |
|  | N | % |  | N | % |  | N | % |
| 2 | 4 | 5.5 | <30 min | 47 | 64.4 | >1 and<br>≤2h | 2 | 2.7 |
| 3 | 28 | 38.4 | 30-39<br>min | 23 | 31.5 | >2 and<br>≤3h | 6 | 8.2 |
| 4 | 33 | 45.2 | 40-49<br>min | 3 | 4.1 | >3 and<br>≤4h | 30 | 41.1 |
| 5 | 5 | 6.9 | Total | 73 | 100 | >4 and<br>≤5h | 26 | 35.6 |
| 6 | 2 | 2.7 |  |  |  | >5 and<br>≤6h | 6 | 8.2 |
| 7 | 1 | 1.4 |  |  |  | >6 and<br>≤7h | 3 | 4.1 |
| Total | 73 | 100 |  |  |  | Total | 73 | 100 |

Most matched applicants (52/73, 71%) felt they learned about programs somewhat (47, 64%) or much (5, 7%) less well than had recruitment been in-person; 20 (27%) felt they learned about programs equally well, and 1 (1%) somewhat better. Despite this perspective, most (56/73, 77%) applicants want to at least have the option for virtual recruitment moving forward, with 32 (44%) preferring face-to-face with an option for virtual, 15 (21%) preferring virtual with an option for face-to-face, and 9 (12%) preferring virtual. Only 17 (23%) prefer in-person interviewing with no virtual option.

### Open-Ended Reponses

When asked to describe the aspect(s) of virtual recruitment/interviewing they are most likely to retain moving forward, 47/79 (59%) of PDs provided at least one response. Of these 47, 28 (60%) plan to retain and/or improve or expand their reservoir of pre-recorded online resources. The second most common response, described by 16 (34%), was a plan to retain the option for either primary or secondary virtual visits, such as for additional meetings with faculty or asynchronous conference viewing. Other responses provided more than twice included maintenance of conversations or “happy hours” with current fellows (5, 11%), an emphasis on diversity, inclusion, and/or avoidance of bias (3, 6%), and a modified interview day structure or format (3, 6%).

Of the 38 PDs who provided at least one aspect of recruitment they are likely to change, 14 (37%) plan to augment or improve their portfolio of pre-recorded online resources. The second most common response was to make no changes (7 [18%]). Other responses provided more than twice included a return to in-person recruitment for at least some component (3, 8%) and inclusion of applicants in conferences if virtual recruitment persists (3, 8%).

When asked to describe the aspect(s) of virtual recruitment/interviewing they would most like to see retained moving forward, 37/85 (44%) matched applicants responded. More than half (20/37, 54%) would like programs to retain their expanded online content. The second most common response, described by 11 (30%), was a preference to retain virtual interviews, whether for primary or secondary visits. Other responses provided more than twice included shorter interview days and/or interview duration (5, 14%) and spending time with fellows, including meetings with smaller groups of fellows (3, 8%).

Of the 28 matched applicants who described aspect(s) of virtual recruitment/interviewing they would most like to see changed moving forward, 15 (54%) indicated a preference for different aspects of the timing or structure of the interview day, including 6 (21%) who preferred shorter interview days, 3 (11%) limits upon the total number of interviews and/or their duration, 2 (7%) sufficiently long breaks between interviews, 2 (7%) consolidation of all interviews into a single day, 1 (4%) interviews spread over the course of the week, and 1 (4%) time for lunch. There were nine (32%) comments pertaining to time spent with fellows with no consensus message. An interest in being able to spend time with co-applicants without program representatives present was expressed by two (7%) respondents, and another referenced the difficulty meeting co-applicants via virtual recruitment. Three comments referenced the importance of information posted by programs on their websites.

## Discussion

While the majority of surveyed PDs and matched applicants prefer in-person recruitment, most want at least the option of virtual recruitment, and nearly all PDs felt they adequately learned about candidates virtually. Additional attributes of virtual recruitment identified by our study include programs’ and applicants’ abilities to accommodate increased numbers of applications and interviews, the value applicants place on specific aspects of programs’ expanded online profiles, and applicants’ preference for time-limited interview days.

The 2020 ID fellowship Match results were notably improved over recent years. There were 404 applicants to the National Resident Matching Program, up from 335-356 over each of the preceding four years.^30^ Of these 404 applicants, 365 (90%) matched into an ID fellowship, such that 365/414 (88%) offered positions filled, up from 79-81% over the preceding four years.^30^ In concert with these Match results, our survey data suggest ID fellowship programs were able to effectively pivot to virtual recruitment. During meetings of ID fellowship PDs in advance of 2020 recruitment, many expressed an expectation that virtual recruitment would be associated with increased application numbers. Our data support this hypothesis, with programs reporting a greater number of applications, interview offers, and interviews than in the prior year. While the majority of applicants reported applying to the same number of programs as they would have had recruitment been in-person, most who modified this number due to virtual recruitment applied to more programs than they otherwise would have. Although associated with increased faculty time demands, the system clearly appears to have had capacity for these increased numbers, and the majority of programs and applicants seem comfortable with them, as most plan to maintain them next year (programs) or would not have changed them if they could (applicants). With the caveat that our study was not designed to address this question as we surveyed only matched applicants, nationally applicants do not appear to have been disadvantaged by the increased number of applicants and applications, as a higher proportion of applicants matched than in recent years, and only 10% of applicants went unmatched, as compared with 7-8% over each of the prior four years.^30^

In concert with previous reports, our data suggest we are unlikely to return to the former all-in-person recruitment status quo.^24,25^ PDs are urged to consider how they will accommodate a mixture of in-person and virtual recruitment once the former again becomes possible, including ways in which they will guard against potential biases towards applicants who choose one format over the other. Our data also provide important perspectives on recruitment preparations. While all online resources were helpful to some proportion of matched applicants, some were rated as critically or very important by at least 60% of applicants, including prose- and photo-based content on program structure, PD program overview video, presentations/testimonials from fellows, prose- and photo-based content on didactics and curriculum, and profiles of current fellows. Programs planning to revise, update, or newly create web content may wish to focus their efforts on these highest-yield areas. When undertaking these preparations, programs should be cognizant of the substantial time investment required to prepare for virtual recruitment, with half of programs anticipating a need for similar resources should virtual recruitment be incorporated into the next recruitment season. These data may help PDs who aim to maintain or increase the support they receive for their PD work, especially in light of the increased number of applicants in 2020 and the expectation that these numbers will be maintained should virtual recruitment continue.

Matched applicants expressed clear preferences regarding the interview day duration and structure. Most prefer 3-4 interviews lasting no more than 40 minutes each, and a total interview day duration of 3-5 hours. We did not solicit input from PDs on these items, but there is a notable difference in the frequency with which these topics were discussed in PD and matched applicant open-ended responses regarding innovation opportunities for the future. PDs are encouraged to be mindful of this input as they plan future recruitment seasons.

Limitations of our study include our PD response rate of 43%. While incomplete, this response rate to an unincentivized survey in the midst of the ongoing COVID-19 pandemic was greater than we anticipated. Although our matched applicant response rate was lower, we are not able to quantify this rate with certainty as the total number of matched applicants who were sent the survey is not known. PD reports of the additional faculty and staff time spent on recruitment activities in 2020 were likely best estimates. The majority of matched applicants only go through the ID fellowship match once and never went through an in-person ID fellowship match; their estimated number of applications and interviews had recruitment been in-person may be different from what would have transpired with an all-in-person recruitment season. We do not have data on the extent to which virtual recruitment permitted applicants with more limited resources to match at programs to which travel costs might have been prohibitive had recruitment been undertaken in-person. Drawing conclusions from free-text responses to survey questions can be misleading, and formal qualitative approaches to analysis of such responses is often inappropriate.^29^ Because these responses complemented the data from responses to Likert-scale style questions, however, we have reported those results through a quantitative content analysis approach. We are unable to exclude the possibility of recall bias and unintentional data entry errors by participants.

Future research efforts should study the perceptions and ramifications of hybrid in-person and virtual recruitment seasons, including strategies for mitigating bias for or against applicants who choose one interview format over the other, as well as cost considerations for applicants and programs. Also important will be to study whether application numbers per program will continue to increase, and how programs balance interview capacity with demand.

## Conclusions

Our survey data and the Match results indicate that the 2020 ID fellowship virtual recruitment season was a remarkable success. With virtual recruitment likely here to stay in some capacity for all specialties and programs, applicants’ views regarding essential aspects of programs’ online portfolios and their preference for time-limited interview days should help inform future recruitment efforts to the benefit of programs and applicants alike.

## Data Availability

All survey responses were transmitted into a password-protected file within the Johns Hopkins University School of Medicine Qualtrics account.

## Notes

### Competing Interest Statement

The authors have declared no competing interest.

### Funding Statement

No external funding was received to support this work.

### Author Declarations

This study was acknowledged as exempt by the Johns Hopkins University School of Medicine Institutional Review Board.

