## Supplemental Appendix 1 for "Virtual Recruitment is Here to Stay: 2020 ID Fellowship Program and Matched Applicant Recruitment Experiences"

**Program Director Survey**

How many applications were submitted to your program in 2020?

[ ]

How many applications were submitted to your program in 2019?

[ ]

How many applicants did you invite for interviews in 2020?

[ ]

How many applicants did you invite for interviews in 2019?

[ ]

How many applicants did you actually interview in 2020?

[ ]

How many applicants did you actually interview in 2019?

[ ]

If recruitment remains virtual next year, what number of applicants do you anticipate interviewing?

- ○ Fewer applicants than in 2020
- ○ Same number as in 2020
- ○ More applicants than in 2020

Did you either post on your program's external website as of 8/12/20 (the date on which applications became available to PDs) or make available to applicants invited to interview the following content?

| | Yes; we newly created this content for 2020 recruitment | Yes; we modified previously-created content for 2020 recruitment | Yes; we maintained previously-created content without change | No, we did not incorporate this content |
|---|---|---|---|---|
| Professionally- or program-made program overview video | ○ | ○ | ○ | ○ |
| Program overview video by PD covering program nuts and bolts | ○ | ○ | ○ | ○ |
| Presentations/testimonials from faculty other than PD | ○ | ○ | ○ | ○ |
| Presentations/testimonials from fellows | ○ | ○ | ○ | ○ |
| Presentations/testimonials from program alumni | ○ | ○ | ○ | ○ |
| Live, virtual attendance at departmental/division conference(s) | ○ | ○ | ○ | ○ |
| Links to departmental/division conference(s) to permit asynchronous attendance | ○ | ○ | ○ | ○ |
| Profiles of current fellows | ○ | ○ | ○ | ○ |
| List of alumni and where they went after fellowship graduation | ○ | ○ | ○ | ○ |
| Prose and photo-based content on program structure, training sites, clinical rotations, and research requirements | ○ | ○ | ○ | ○ |
| Prose and photo-based content on didactics and curriculum | ○ | ○ | ○ | ○ |
| Links to content from institutional GME office | ○ | ○ | ○ | ○ |
| Links to virtual tour(s) of your hospitals, clinic sites, university, campus, and/or clinical setting | ○ | ○ | ○ | ○ |

| | Yes; we newly created this content for 2020 recruitment | Yes; we modified previously-created content for 2020 recruitment | Yes; we maintained previously-created content without change | No, we did not incorporate this content |
|---|---|---|---|---|
| Instructions for technological aspects of the interview season (e.g., description of format, computer/laptop, internet access, virtual platform, or other information) | ○ | ○ | ○ | ○ |
| Other (please describe) ____ | ○ | ○ | ○ | ○ |

Did transitioning to a virtual interview format require any additional resources, whether faculty time, staff time, or funds, as compared to the in-person format?

- ○ Yes
- ○ No

How much additional PD + APD time was required (e.g., planning meetings, website content creation/development, interviewing applicants) beyond time usually spent during recruitment?

- ○ No additional PD + APD time was required
- ○ <10 hours
- ○ 10-19 hours
- ○ 20-29 hours
- ○ 30-39 hours
- ○ 40-49 hours
- ○ ____ >50 hours (enter estimated number of hours)

How much additional total faculty time (other than PD + APD) was required (e.g., videos for website, interviewing applicants) beyond time usually spent during recruitment?

- ○ No additional faculty (other than PD + APD) time was required
- ○ <10 hours

- 10-19 hours
- 20-29 hours
- 30-39 hours
- 40-49 hours
- [ ] >50 hours (enter estimated number of hours)

How much additional total staff/personnel time was required (e.g., virtual platform training, communications with PD/APD, communications with other faculty) beyond time usually spent during recruitment?

- No additional staff/personnel time was required
- <10 hours
- 10-19 hours
- 20-29 hours
- 30-39 hours
- 40-49 hours
- [ ] >50 hours (enter estimated number of hours)

What was the additional monetary cost of virtual recruitment (e.g., videography, webcams, new or updated videoconferencing technology, gifts) beyond funds usually spent during recruitment?

- No additional monetary costs were incurred
- <$100
- $100-$249
- $250-$499
- $500-$999
- [ ] >$1000 (enter estimated additional cost)

If you encountered additional costs not covered by the preceding questions, please describe them here:

Now that your program has conducted a virtual recruitment season, if next year's recruitment season is conducted in a virtual format, how would you estimate the resources needed (time, funding, personnel) would compare to this year?

- Fewer resources will be needed
- Similar resources will be needed
- More resources will be needed

Did you feel that you were able to learn about each candidate sufficiently through the virtual recruitment process?

- Yes, even more so than during face-to-face recruitment
- Yes, equally well as during face-to-face recruitment
- Yes, but not as well as during face-to-face recruitment
- No

Did you feel that you were able to adequately showcase your program through virtual recruitment?

- Yes, even more so than during face-to-face recruitment
- Yes, equally well as during face-to-face recruitment
- Yes, but not as well as during face-to-face recruitment
- No

Assuming resolution of the pandemic, would you prefer recruitment be virtual or in-person for future years? Consider only the primary interview day (e.g., not a "second look").

- In-person
- In-person as default with option to convert to virtual
- Virtual as default with option to convert to in-person

○ Virtual

What aspect(s) of virtual recruitment/interviewing are you most likely to retain moving forward, regardless of whether virtual recruitment continues?

What aspect(s) of virtual recruitment/interviewing are you most likely to change moving forward, regardless of whether virtual recruitment continues?

Powered by Qualtrics
