## Supplemental Appendix 2 for "Virtual Recruitment is Here to Stay: 2020 ID Fellowship Program and Matched Applicant Recruitment Experiences"

**Matched Applicant Survey**

To how many programs did you apply in 2020?

[ ]

Was this number influenced by the virtual nature of the 2020 interview season?

- ○ Yes
- ○ No

In what way did the virtual format affect the number of programs to which you applied?

- ○ I applied to fewer programs than I would have had interviews been in-person
- ○ I applied to 1-3 additional programs than I would have had interviews been in-person
- ○ I applied to 4-6 additional programs than I would have had interviews been in-person
- ○ I applied to >6 additional programs than I would have had interviews been in-person

How many interview offers did you receive in 2020?

[ ]

How many interview days did you actually attend in 2020?

[ ]

Was this number influenced by the virtual nature of the 2020 interview season?

- ○ Yes
- ○ No

In what way did the virtual format affect the number of programs at which you interviewed?

- ○ I applied to fewer programs than I would have had interviews been in-person
- ○ I applied to 1-3 additional programs than I would have had interviews been in-person
- ○ I applied to 4-6 additional programs than I would have had interviews been in-person
- ○ I applied to >6 additional programs than I would have had interviews been in-person

If you could go back and do it again, to how many programs would you have applied in 2020?

- More
- The same number
- Fewer

If you could go back and do it again, at how many programs at which you were offered an interview would you have interviewed in 2020?

- More
- The same number
- Fewer

In deciding whether to apply to a program, in helping you prepare for your interview day, and/or in helping you craft your rank list, how helpful to you were each of the following features of programs' websites?

| | Critically important; essential | Very important; not essential, but would not have wanted to miss | Modestly important; helped round out the edges | Neutral importance; interesting but did not impact my view of the program | Not important; not needed |
|---|---|---|---|---|---|
| Professionally- or program-made program overview video | O | O | O | O | O |
| Program overview video by PD covering program nuts and bolts | O | O | O | O | O |
| Presentations/testimonials from faculty other than PD | O | O | O | O | O |
| Presentations/testimonials from fellows | O | O | O | O | O |
| Presentations/testimonials from program alumni | O | O | O | O | O |
| Live, virtual attendance at departmental/division conference(s) | O | O | O | O | O |

| | Critically important; essential | Very important; not essential, but would not have wanted to miss | Modestly important; helped round out the edges | Neutral importance; interesting but did not impact my view of the program | Not important; not needed |
|---|---|---|---|---|---|
| Links to departmental/division conference(s) to permit asynchronous attendance | ○ | ○ | ○ | ○ | ○ |
| Profiles of current fellows | ○ | ○ | ○ | ○ | ○ |
| List of alumni and where they went after fellowship graduation | ○ | ○ | ○ | ○ | ○ |
| Prose and photo-based content on program structure, training sites, clinical rotations, and research requirements | ○ | ○ | ○ | ○ | ○ |
| Prose and photo-based content on didactics and curriculum | ○ | ○ | ○ | ○ | ○ |
| Links to content from institutional GME office | ○ | ○ | ○ | ○ | ○ |
| Links to virtual tour(s) of hospital, clinic site, university, campus, and/or clinical setting | ○ | ○ | ○ | ○ | ○ |
| Interview day virtual platform-based group discussion among fellows and applicants | ○ | ○ | ○ | ○ | ○ |
| Pre- or post-interview day virtual platform-based group discussion among fellows and applicants | ○ | ○ | ○ | ○ | ○ |
| Instructions for technological aspects of the interview day (e.g., description of format, computer/laptop, internet access, virtual platform, or other information) | ○ | ○ | ○ | ○ | ○ |
| Other (please describe) ______ | ○ | ○ | ○ | ○ | ○ |

In your opinion, what number of faculty interviews on a given day would achieve the ideal balance between meeting different faculty and learning about a program while avoiding interview fatigue?

- ○ 1
- ○ 2
- ○ 3
- ○ 4
- ○ 5
- ○ 6
- ○ 7
- ○ 8
- ○ 9
- ○ >=10

In your opinion, what faculty interview duration would achieve the ideal balance between getting an interviewer's perspectives on the program and letting them get to know you while avoiding interview fatigue?

- ○ <30 minutes
- ○ 30-39 minutes
- ○ 40-49 minutes
- ○ 50-59 minutes
- ○ >=60 minutes

What is your preference for the structure of an interview day?

- ○ Consolidated into one continuous block of time on a single day
- ○ Spread over different times on a single day
- ○ Spread over different days
- ○ ________________ Other (please describe)

In your opinion, what is the ideal duration for breaks between individual interviews?

- ○ I prefer no break between interviews

- <5 minutes
- 5-9 minutes
- 10-14 minutes
- 15-19 minutes
- >=20 minutes

If a program sent you a gift box, voucher/coupon for food, or comparable item, how did you view this item and incorporate its receipt into your ranking decision?

- Very favorably; I ranked at least one program higher specifically because of this item
- Favorably, though I don't think I ranked any program higher as a direct result
- Neutral
- Unfavorably, though I don't think I ranked any program lower as a direct result
- Very unfavorably; I ranked at least one program lower specifically because of this item

In your opinion, how long should the virtual interview day last overall in order to achieve a balance between learning about a program and avoiding interview fatigue?

- Up to one hour
- More than 1 and up to 2 hours
- More than 2 and up to 3 hours
- More than 3 and up to 4 hours
- More than 4 and up to 5 hours
- More than 5 and up to 6 hours
- More than 6 and up to 7 hours
- More than 7 and up to 8 hours
- More than 8 hours

To what extent do you feel you were able to adequately learn about programs through virtual recruitment?

- Much better than if recruitment had been in-person
- Somewhat better than if recruitment had been in-person
- Equally well as if recruitment had been in-person
- Somewhat less well than if recruitment had been in-person

- Much less well than if recruitment had been in-person

Assuming resolution of the pandemic, would you prefer recruitment be virtual or in-person for future years? Consider only the primary interview day (e.g., not a "second look").

- In-person
- In-person as default with option to convert to virtual
- Virtual as default with option to convert to in-person
- Virtual

What aspect(s) of virtual recruitment/interviewing would you most like to see retained moving forward, regardless of whether virtual recruitment continues?

What aspect(s) of virtual recruitment/interviewing would you most like to see changed moving forward, regardless of whether virtual recruitment continues?

Powered by Qualtrics
